# Performance of COVIDSeq and Swift normalase amplicon SARS-CoV-2 panels for SARS-CoV-2 Genomes Sequencing: Practical Guide and Combining FASTQ Strategy

**DOI:** 10.1101/2021.12.30.21268553

**Authors:** Hosoon Choi, Munok Hwang, Dhammika H. Navarathna, Jing Xu, Janell Lukey, Chetan Jinadatha

## Abstract

The whole genomic sequencing (WGS) of SARS-CoV-2 has been performed extensively and is playing a crucial role in fighting against COVID-19 pandemic. Obtaining sufficient WGS data from clinical samples is often challenging especially from the samples with low viral load. We evaluated two SARS-CoV-2 sequencing protocols for their efficiency/accuracy and limitations. Sequence coverage of >95% was obtained by Swift normalase amplicon SARS-CoV-2 panels (SNAP) protocol for all the samples with Ct ≤ 35 and by COVIDSeq protocol for 97% of samples with Ct ≤ 30. Sample RNA quantitation obtained using digital PCR provided more precise cutoff values. The quantitative digital PCR cutoff values for obtaining 95% coverage are 10.5 copies/μL for SNAP protocol and 147 copies/μL for COVIDSeq protocol. Combining FASTQ files obtained from 2 protocols improved the outcome of sequence analysis by compensating for missing amplicon regions. This process resulted in an increase of sequencing coverage and lineage call precision.

## INTRODUCTION

The pandemic coronavirus disease-2019 (COVID-19) caused by severe acute respiratory syndrome coronavirus 2 (SARS-CoV-2) brought dire consequences and affected not only the physical health of individuals but also caused serious socioeconomic devastation (1, 2). The complete genomic sequences of SARS-CoV-2 were obtained in late December 2019 (3-5) and played crucial role in developing detection methods including real-time PCR based diagnostic tests which became the gold standard for COVID-19 test (6). Real-time RT-PCR diagnostic tests were able to detect the SARS-CoV-2 even from the asymptomatic or mildly symptomatic patients which contributed to appropriate quarantine hence preventing transmission of COVID-19 within the community.

Along with the diagnostic testing, whole-genome sequencing (WGS) of SARS-CoV-2 has been an irreplaceable tool for investigating the evolution of the SARS-CoV-2 and its impact on the disease severity and global spread (7, 8). Genomic sequence analysis also has provided essential information in the development of antiviral therapeutics and vaccines (9, 10). With substantial progress and expansion of the sequencing techniques, WGS of SARS-CoV-2 can be readily performed in many research labs and provides crucial information for thorough epidemiological analysis (11, 12). WGS is an important tool to reveal epidemiological relatedness of transmission and detection of mutation for potential variants of concern. Many researchers worldwide developed bioinformatic SARS-CoV-2 genome analysis tools such as Pangolin lineage (https://pangolin.cog-uk.io/) and Nextclade (https://clades.nextstrain.org/). Due to these efforts, since the first appearance of Wuhan virus, numerous SARS-CoV-2 variants, such as Alpha, Beta, Gamma, Delta, and Omicron, along with its significant mutations in SARS-CoV-2 genome were discovered through WGS.

The limitation of WGS for SARS-CoV-2 is that it needs high viral load to get enough sequencing coverage for obtaining information such as lineage, clade, or mutations. For example, at least 50% of genome must be good reads without N bases call to obtain sample lineage at Pangolin. The accuracy of lineage call depends on the higher sequencing coverage. Therefore, low viral load samples are often not subject to WGS in many labs. However, considering substantial number of vaccinated individuals who become infected and are either asymptomatic or mildly symptomatic causing breakthrough infections to be less likely detected (13). WGS of low viral load patient samples is necessary to detect and break transmission chains involving immune-escape variants.

Based on the analysis of 2 amplicon-based sequencing methods, COVIDSeq (Illumina) and Swift normalase amplicon SARS-CoV-2 panels (SNAP) (Swift Biosciences) (14-16), here we report WGS guide for SARS-CoV2 including low viral load samples. Through testing of various viral load samples with amplicon-based library prep kits along with digital PCR, we were able to obtain tentative cutoff value of sample viral load above which one can expect to acquire informative sequencing outcome. In addition, combining FASTQ strategy has been applied to maximize the sequencing coverage and obtain sequence information even from samples with low viral load. The strategy to combine FASTQ files could also be effective in case of amplicon loss (17) that could occur due to novel mutations in emerging variants like Omicron.

## MATERIALS AND METHODS

### RNA extraction

RNAs were isolated from de-identified clinical samples (nasopharyngeal swab) diagnosed as COVID-19 positive in Central Texas Veterans Health Care System, Temple, TX. RNAs were extracted using QIAmp Viral RNA Kit (Qiagen, Hilden, Germany, catalog number. 529006) according to the manufacture’s protocol. For samples with high Ct value (>30), RNA was concentrated with increased sample volume (280 μL) and reduced elution volume (30 μL). Sample Ct value was obtained by direct RT-qPCR using the BD MAX™ SARS-CoV-2 assay (BD, Franklin Lakes, NJ).

### RNA quantification by digital PCR

For quantification of viral load in extracted RNA, digital PCR analysis were performed using TaqPath COVID-19 combo kit (Thermo Fisher Scientific, Waltham, MA, USA, catalog number A47814). Digital PCR analysis with extracted RNA was performed with QIAcuity (Qiagen) with QIAcuity Probe PCR kit 2 step prep (Qiagen Cat. No 250101) with 26K partition per well nanoplates (Qiagen Cat. No 250001) according to the manufacturer’s protocol. 5 μL of cDNA synthesized with COVIDseq Test kit was used in 40 μL reaction volume.

### Library preparation and sequencing

From the extracted RNA, libraries for sequencing were prepped using COVIDseq Test (Illumina, San Diego, CA, USA, catalog number 20043675). and Swift normalase amplicon SARS-CoV-2 panels (SNAP) library prep kit (Swift biosciences, Ann Arbor, MI, USA, catalog number SN-5X296) according to the manufacture’s protocol. The cDNAs synthesized with COVIDseq Test kit were used as starting materials for Swift library prep. Optional normalase I and normalase II treatment was performed on all samples. For both kit, maximum recommended number of PCR cycles was applied. Sequencing of prepared libraries was performed using the Illumina 500/550 Mid Output Kit (Illumina, catalog number 20024905) and NextSeq 550 system according to manufacturer’s protocol with paired-end reads (2 × 149 bp). 1.4pM of pooled library with 1% phiX was loaded.

### Combining FASTQ strategy & data analysis

After sequencing run, NextSeq Local Run manager was used to generate FASTQ files. Illumina BaseSpace Sequence Hub was used for further analysis by uploading FASTQ filles. FASTQ files were assembled through reference mapping against SARS-CoV-2 sequence (NC_045512) using Illumina SARS-CoV-2 NGS Data Toolkit, DRAGEN COVID Lineage App and generated consensus FASTA files. Consensus FASTA files were generated that meet criteria of variant frequency (VF) ≥ 0.5 and coverage >10x. If VF<0.5 and coverage >10x, the base marked as ‘N’. Genome coverage with percent of non-N bases and median coverage (read depth) was calculated. Consensus FASTA files were used to identify the lineage and clade of samples using Pangolin lineage (https://pangolin.cog-uk.io/) and Nextclade (https://clades.nextstrain.org/), respectively. For samples whose lineage call is different or poor sequencing coverage from COVIDseq and Swift prep, FASTQ files from both library preps were combined by uploading files under the same Biosample at BaseSpace. For detailed sequence analysis, NCBI BLAST (https://blast.ncbi.nlm.nih.gov/Blast.cgi) was used.

## RESULTS

A total of 121 samples from patients with COVID-19 positive RT-qPCR test were subject to WGS and quantitative digital PCR. Two parameters were used for evaluating SARS-CoV-2 WGS outcome. One is the percent genome coverage which indicates good base calls for non-N bases. The other is the median coverage which indicates read depth. Generally, percent genome coverage over 95% with more than 100x median coverage is considered good sequencing coverage.

### Quantification

The initial positive samples’ Ct values obtained from direct RT-qPCR of SARS-CoV-2 N2 gene using BD MAX™ SARS-CoV-2 assay were between 11 and 45. The Ct value of about one third of samples (42 out of 121) were higher than 30 which indicates low viral load. To obtain actual quantity of SARS-CoV-2 genome in isolated RNA, digital PCR were performed using multiplex assay kit (Qiagen Probe PCR kit) for three SARS-CoV-2 genes, N, S, and ORF1ab. As shown in Figure 1A, Ct values obtained from real-time RT-PCR and N gene copy numbers from digital PCR are highly correlated. Moreover, quantitative digital PCR provided precise cutoff values for sequencing outcome. For using SNAP protocol, all the samples with more than 10.7 copies/μL N gene of SARS-CoV-2 produced >95% genome coverage (Figure 2C). Only 6 out of 20 samples with <10.5 copies/μL produced more than 95% coverage (Figure 3C). With COVIDSeq test kit, all the samples with more than 147 copies/μL produced >95% genome coverage (Figure 2D) while only 5 samples less than 147 copies/μL yielded producing >95% coverage (Figure 3D).

**Figure 1.**
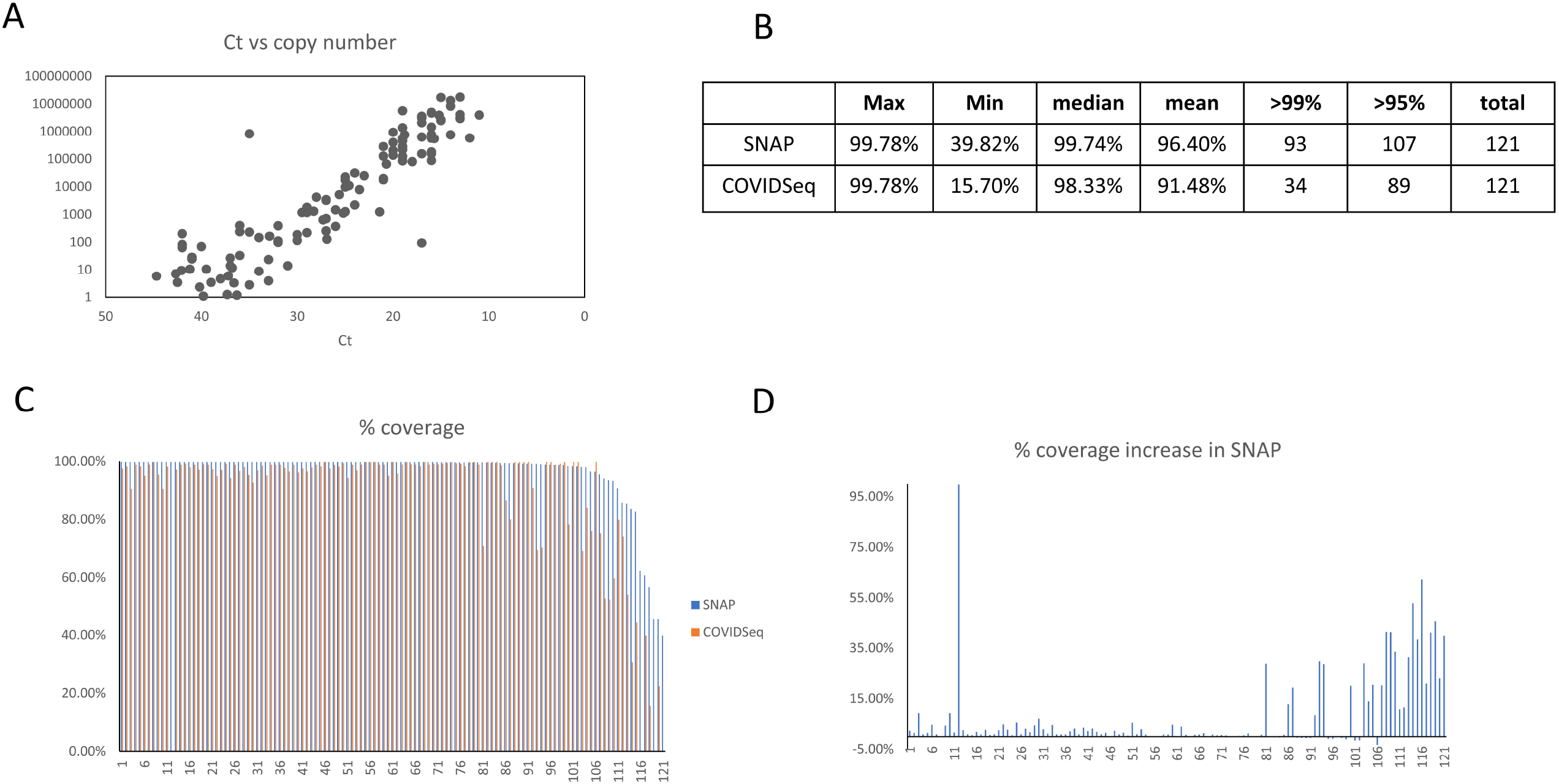
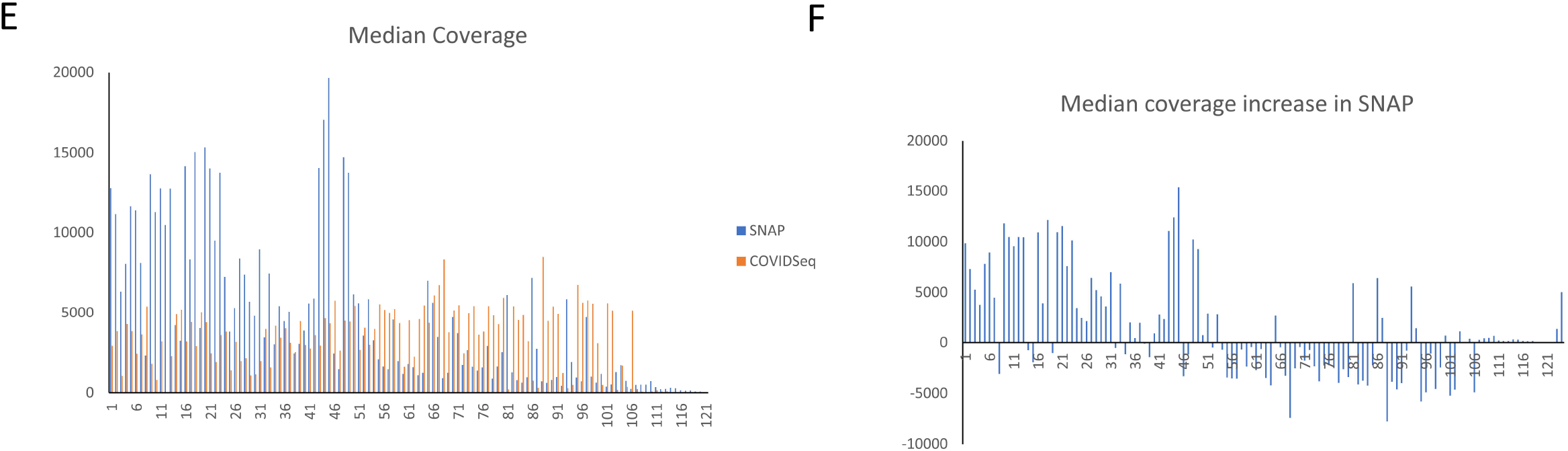
Overview of the SARS-CoV-2 sequencing outcome of SNAP protocol and COVIDSeq protocol. (A) Correlations between Ct values and copy numbers obtained using digital PCR. (B) Summary of sequencing coverage of SNAP protocol and COVIDSeq protocol. (C) % coverage of SNAP protocol and COVIDSeq protocol for each sample sequenced. (D) % coverage increases in SNAP protocol compare to COVIDSeq protocol. (E) Median coverage of SNAP protocol and COVIDSeq protocol for each sample sequenced. (D) Median coverage increases in SNAP protocol compare to COVIDSeq protocol.

**Figure 2.**
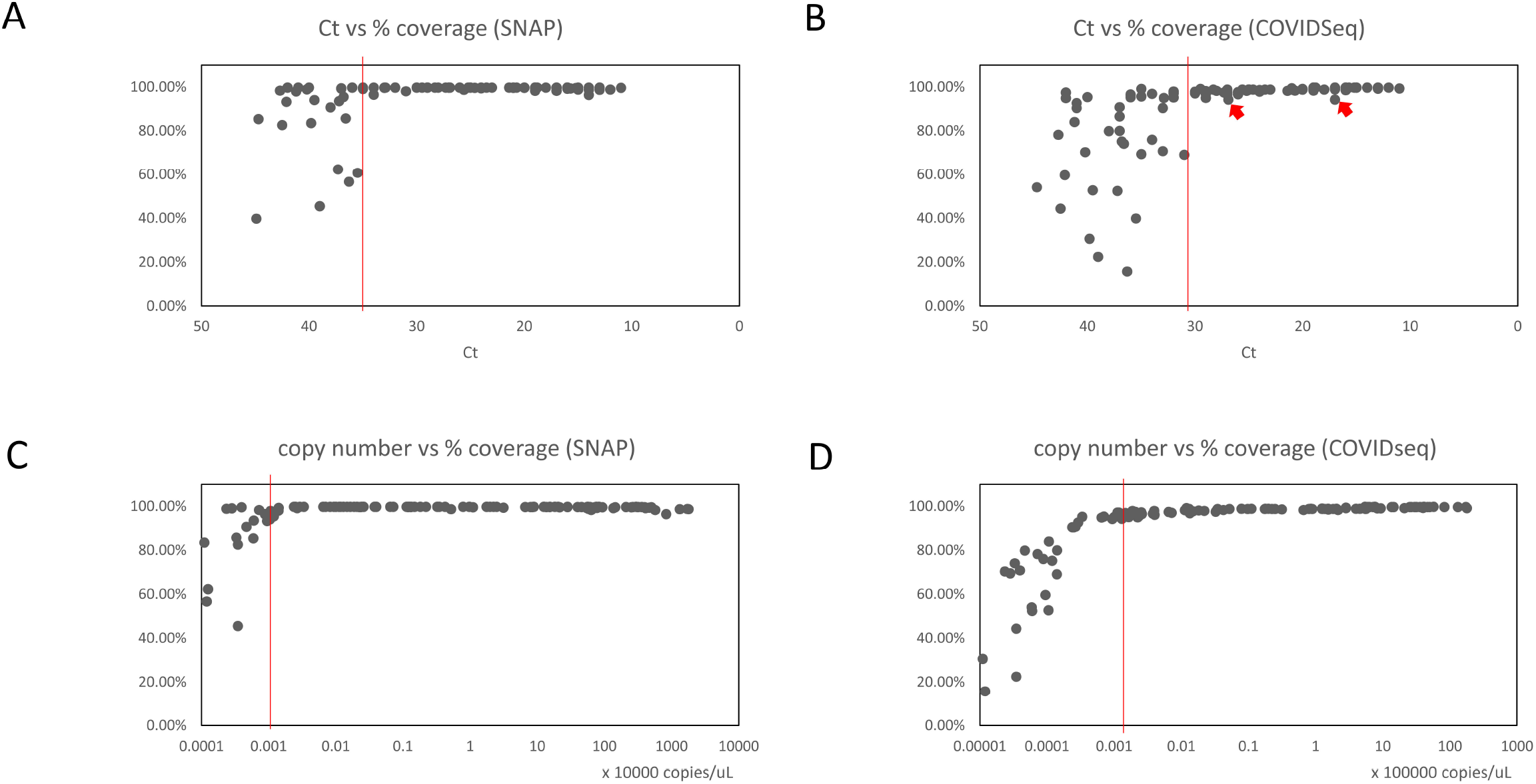
Correlation between viral loads and % coverage. (A) Correlations between Ct values and % coverage of SNAP protocol. (B) Correlations between Ct values and % coverage of COVIDSeq protocol. (C) Correlations between copy number obtained by digital PCR and % coverage of SNAP protocol. (B) Correlations between copy number obtained by digital PCR and % coverage of COVIDSeq protocol. Red bars are proposed Ct cutoff with which all the samples located to right side produced >95% genome coverage. Two exceptions in B are indicated by arrows.

**Figure 3.**
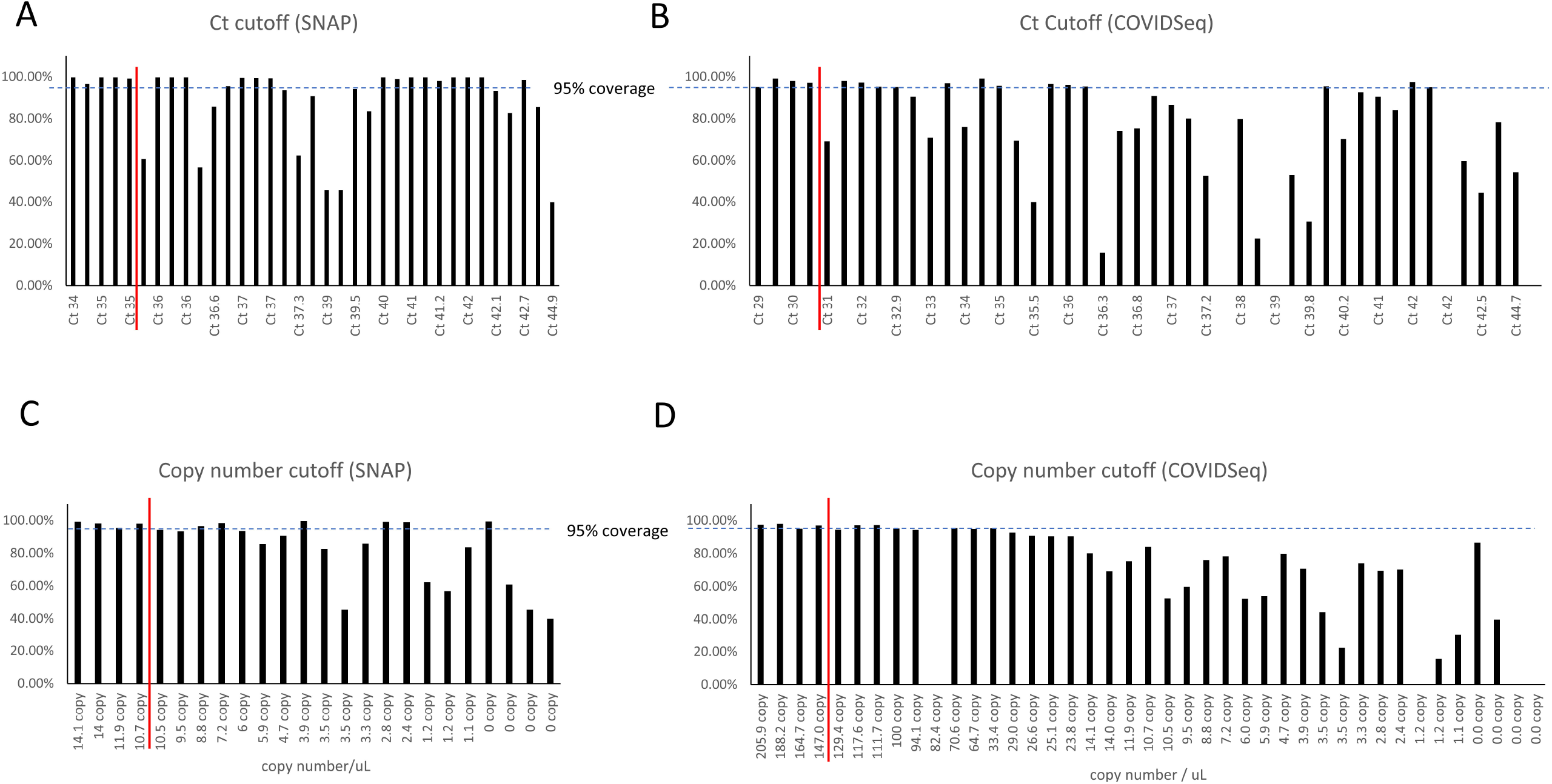
% coverage of low viral load samples. (A) Distribution of % coverage by SNAP protocol of high Ct samples (B) Distribution of % coverage by COVIDSeq protocol of high Ct samples. (C) Distribution of % coverage by SNAP protocol of low copy samples. (D) Distribution of % coverage by COVIDSeq protocol of low copy samples. Red bars are proposed Ct cutoff with which all the samples located to left side produced >95% genome coverage.

### WGS sequencing

Libraries for SARS-CoV-2 WGS were prepared using amplicon-based library prep kits; SNAP protocol (Swift) and COVIDSeq test (RUO) protocol (Illumina). For each sequencing run, over 85% of base read showed quality score ≥ Q30. Even though one third of the samples were low viral load samples (Ct >30) and some samples’ Ct was over 40, all 121 samples were sequenced with WGS. The average of percent genome coverage for SNAP protocol and COVIDSeq protocol was 96.40% and 91.48%, respectively. Out of 121 samples sequenced, greater than 95% genome coverage was obtained for 107 samples by SNAP protocol and 89 samples by COVIDSeq protocol. The sequence yield from both protocols exceeds many previous SARS-CoV-2 sequencing reports, especially for low viral load samples (Figure 1B).

### Sequencing coverage

Overall, SNAP protocol produced better sequencing coverage with higher % coverage (paired t-test, p=3.2 × 10^−8^) (Figure 1C, 1D) and read depth than COVIDSeq protocol: median coverage of 4579.9 vs 3333.3, paired t-test, p=0.0058 (Figure 1E, 1F). For samples with Ct ≤ 30, both protocols’ yield was similar. SNAP protocols achieved >95% coverage in all samples while COVIDSeq protocol were able to produce >95% coverage from 77 out of 79 samples (Figure 2B). However, in low viral load samples sequencing coverage was better in SNAP. For low viral load samples, while SNAP protocol was able to yield >95% coverage from all 12 samples with Ct = 31∼35 (Figure 2A), only 7 out of 12 samples with COVIDSeq. For samples with Ct >35, SNAP protocol yields > 95% coverage in 16 samples out of 30 samples (Figure 3A) while only 5 samples with COVIDSeq (Figure 3B). Read depth of SNAP protocol ranged from 41X to 19662X median coverage. All samples >750X coverage from SNAP protocol achieved >95% coverage (Figure 4A). COVIDSeq protocol got 3X to 8475X median coverage. Median coverage of >2800X is sufficient to have >95% coverage by COVIDSeq protocol (Figure 4B).

**Figure 4.**
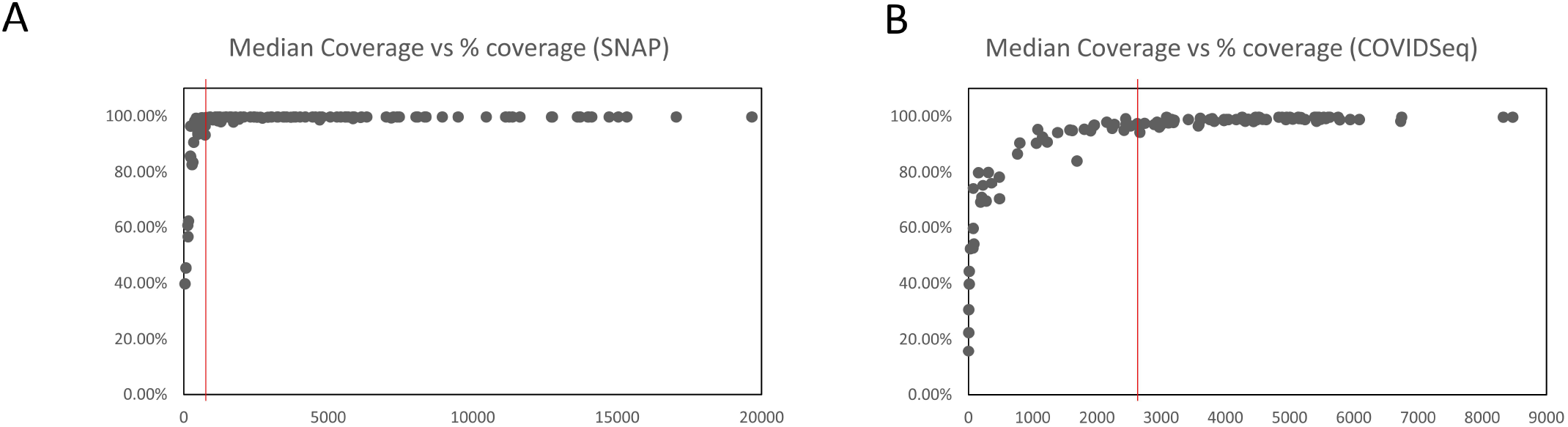
Correlation between median coverage and % coverage. (A) Correlations between median coverage and % coverage of SNAP protocol. (B) Correlations between median coverage and % coverage of COVIDSeq protocol right side of the red bar produced >95% genome coverage.

### Combining FASTQ strategy

To increase sequence reads and coverage, FASTQ files of 52 samples obtained from COVIDSeq and SNAP protocols were combined for analysis. Combining FASTQ improved sequencing coverage significantly with average percent genome coverage 94.58% compare to 92.18% for SNAP and 82.67% COVIDSeq protocol (Figure 5 and 6). 36 out of 52 combined analyzed samples percent coverage increased to >99% coverage compared to 30 samples from SNAP and only 4 samples from COVIDSeq protocol. Combining FASTQ analysis especially impacted the samples with high Ct. Combining FASTQ analysis resulted in median genome coverage of 99.46% for 42 samples with Ct >30. As a result, only 8 samples out of 121 samples’ percent genome coverage were <95% and percent coverage > 95% was obtained for 33 samples with Ct >30 (ranges between Ct 31 and 42.7) by SNAP only or combining COVIDSeq and SNAP (Figure 6C). Sequence depth was increased significantly by combining FASTQ analysis (Figure 6A, 6B, Table 2). However, for the samples with median coverage 400 and below from SNAP protocol, median coverage didn’t reach to the level for reliable lineage calling (COVID lineage app. v 3.5.4).

**Figure 5.**
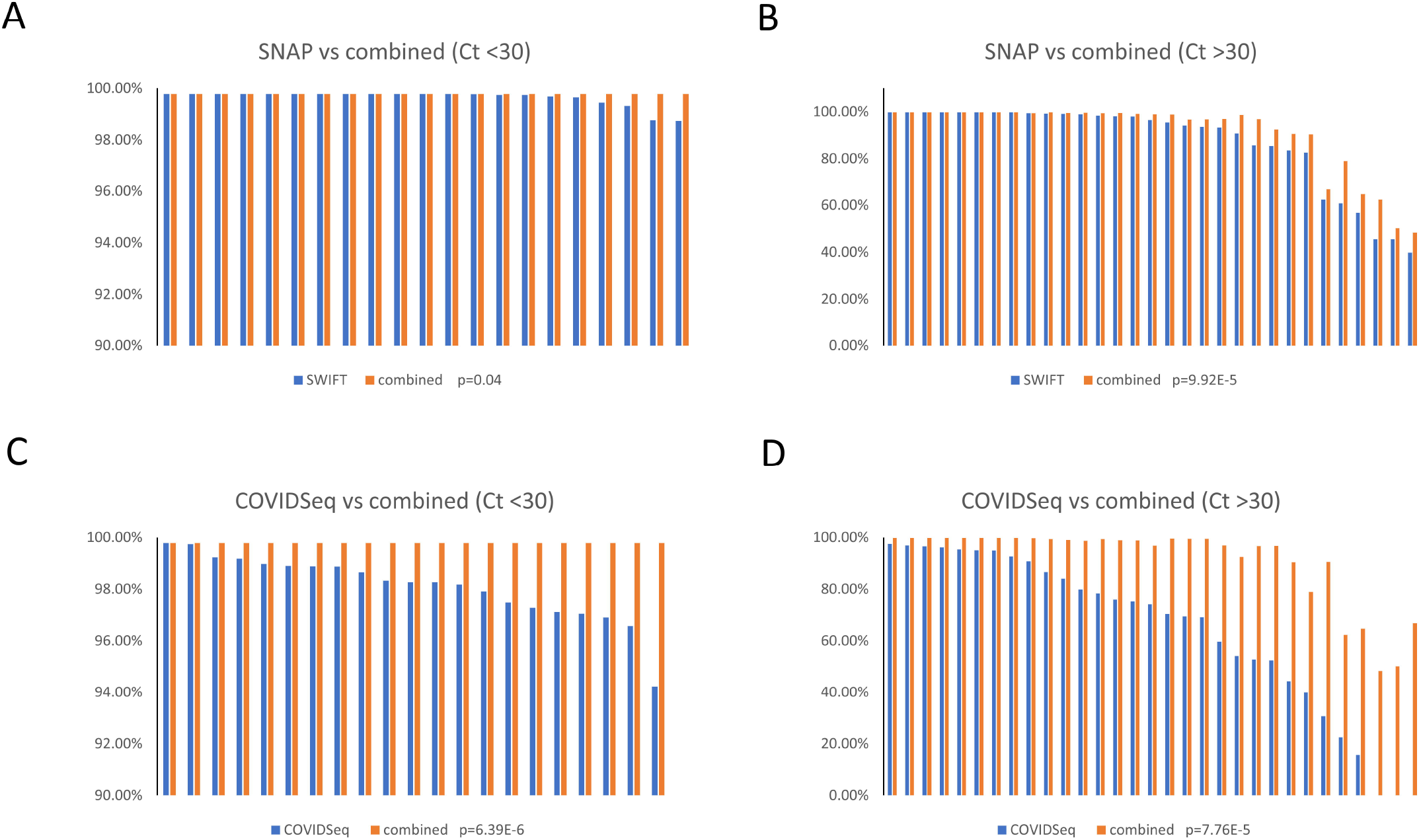
Increases of % coverage by combined FASTQ. (A) Comparison of % coverage between SNAP protocol and combined FASTQ of low CT samples. (B) Comparison of % coverage between SNAP protocol and combined FASTQ of high CT samples. (C) Comparison of % coverage between COVIDSeq protocol and combined FASTQ of low CT samples. (B) Comparison of % coverage between COVIDSeq protocol and combined FASTQ of high CT samples.

**Figure 6.**
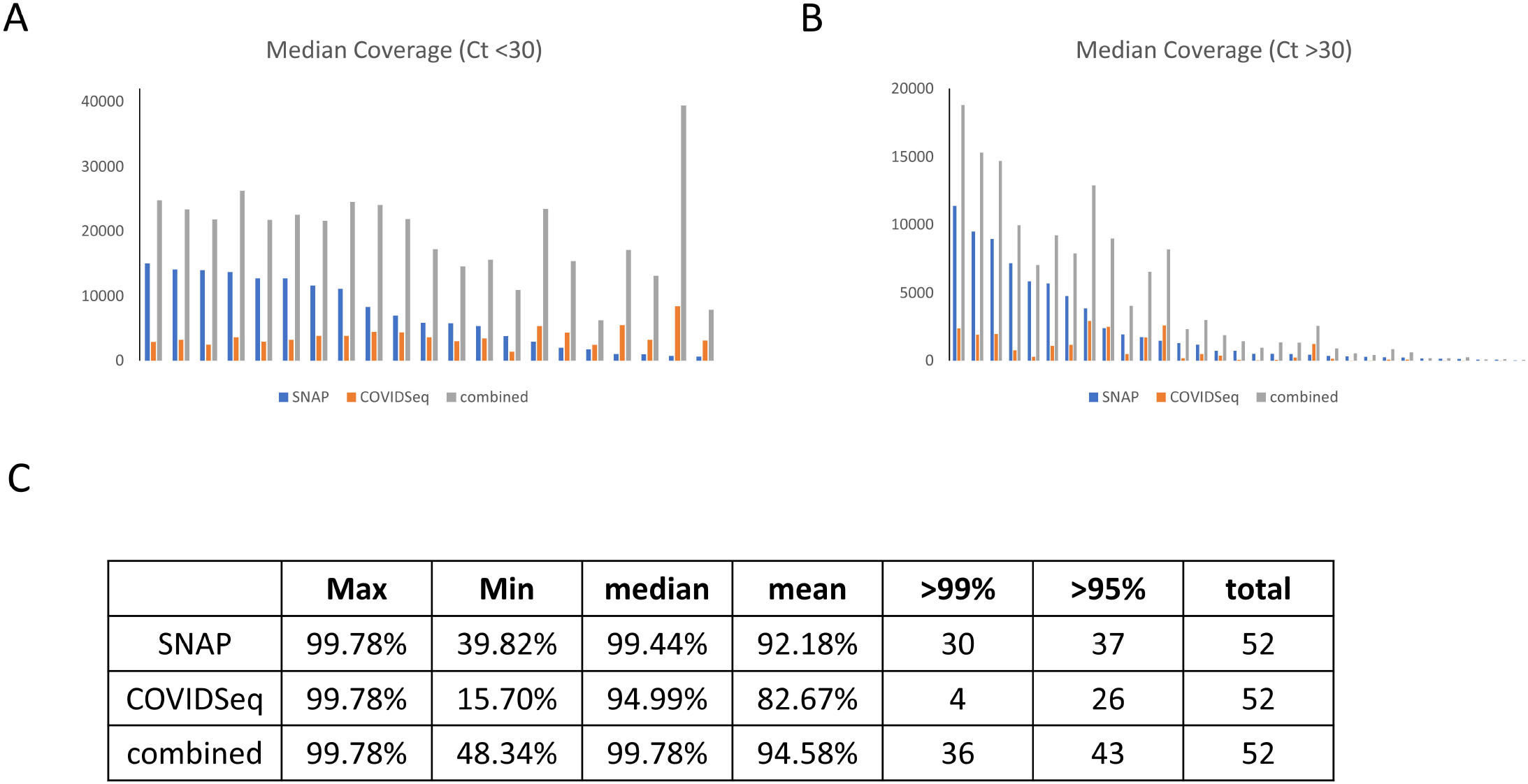
Increases of median coverage by combined FASTQ. (A) Comparison of median coverage low CT samples. (B) Comparison of median coverage high CT samples. (C) Summary of sequencing coverage of SNAP protocol, COVIDSeq protocol, and combined FASTQ.

### Lineage determination

Since the sample collection period overlapped with Delta variants surge in the US, majority of samples (104 out of 121) were Delta (B.1.617.2) or Delta sub-lineages (AY.3∼ AY.26). 16 samples were other SARS-CoV-2 variants, and 1 sample’s (Ct=45) precise lineage was not determined due to the low sequencing coverage. The SARS-CoV-2 lineages call was identical for 69 samples determined by both protocols and were different from each other for 52 samples. Combining FASTQ analysis was performed for samples with lineage call discrepancy. The lineages of 20 out of 52 samples were changed to new lineages that were not same as neither the lineages derived from COVIDSeq or SNAP protocol. For 15 samples, lineage calls of combined FASTQ analysis were agreed with COVIDSeq, and 16 samples agreed with SNAP protocol (Table 1).

**Table 1.** lineage calls derived by SNAP, COVIDSeq, and combined FASTQ.

|  | Lineage (SNAP) | Lineage (COVIDSeq) | Lineage (combined) | Ct VALUE |
| --- | --- | --- | --- | --- |
| PV_13 | AY.4 | B.1.617.2 | AY.5 | 15.7 |
| PV2_63 | AY.10 | **B.1.617.2** | **B.1.617.2** | 14 |
| PV2_39 | B.1.617.2 | **AY.3** | **AY.3** | 13 |
| PV2_62 | B.1.617.2 | **AY.25** | **AY.25** | 17 |
| PV2_93 | **AY.24** | B.1.617.2 | **AY.24** | 19 |
| PV2_49 | B.1.617.2 | **AY.25** | **AY.25** | 20 |
| PV2_55 | AY.24 | **B.1.617.2** | **B.1.617.2** | 21 |
| PV2_30 | AY.12 | **B.1.617.2** | **B.1.617.2** | 25 |
| PV2_40 | AY.25 | AY.12 | **B.1.617.2** | 24 |
| PV2_91 | B.1.617.2 | **AY.5** | **AY.5** | 19 |
| PV2_3 | B.1.617.2 | **AY.3** | **AY.3** | 25.2 |
| PV2_7 | **B.1.617.2** | AY.3 | **B.1.617.2** | 21.4 |
| PV2_18 | **B.1.617.2** | AY.25 | **B.1.617.2** | 29 |
| PV2_23 | AY.10 | B.1.617.2 | **AY.25** | 30 |
| PV2_2 | **B.1.617.2** | AY.12 | **B.1.617.2** | 27.3 |
| PV2_89 | **B.1.617.2** | AY.12 | **B.1.617.2** | 42 |
| PV2_34 | B.1.617.2 | **AY.25** | **AY.25** | 27 |
| PV2_31 | **B.1.617.2** | AY.25 | **B.1.617.2** | 30 |
| PV2_38 | **B.1.617.2** | AY.3 | **B.1.617.2** | 27 |
| PV2_50 | **B.1.617.2** | AY.12 | **B.1.617.2** | 34 |
| PV2_70 | **B.1.617.2** | AY.25 | **B.1.617.2** | 29 |
| PV2_64 | AY.25 | AY.12 | **B.1.617.2** | 36 |
| PV2_76 | **B.1.617.2** | AY.12 | **B.1.617.2** | 27 |
| PV2_67 | **B.1.617.2** | AY.7.1 | **B.1.617.2** | 36 |
| PV2_47 | **B.1.617.2** | AY.25 | **B.1.617.2** | 36 |
| PV2_8 | **AY.3** | AY.12 | **AY.3** | 32.9 |
| PV2_35 | **AY.25** | B.1.617.2 | **AY.25** | 42 |
| PV2_42 | AY.10 | **B.1.617.2** | **B.1.617.2** | 17 |
| PV2_48 | B.1.617.2 | **AY.25** | **AY.25** | 41 |
| PV2_58 | **B.1.617.2** | AY.25 | **B.1.617.2** | 37 |
| PV2_26 | B.1.617.2 | **AY.25** | **AY.25** | 37 |
| PV_12 | B.1 | **P.2** | **P.2** | 41.2 |
| PV_2 | B.1.617.2 | AY.12 | **AY.4** | 38 |
| PV_6 | AY.10 | AY.7.1 | **B.1.617.2** | 42.7 |
| PV2_82 | AY.10 | AY.7.1 | **B.1.617.2** | 34 |
| PV_4 | AY.4 | B.1.575 | **AY.10** | 36.8 |
| PV_1 | B.1 | B.1.1 | **B.1.617.2** | 36.6 |
| PV_11 | AY.10 | B.1.617.2 | **AY.4** | 40.2 |
| PV2_25 | B.1.617.2 | **AY.3** | **AY.3** | 35 |
| PV2_83 | AY.10 | B.1.617.2 | **AY.25** | 31 |
| PV_5 | B.1.2 | **B.1.1.519** | **B.1.1.519** | 42.1 |
| PV_3 | B.1.153 | B.1 | **B.1.617.1** | 44.7 |
| PV_10 | AY.21 | ND | **B.1.617.2** | 39.5 |
| PV_7 | B.1.153 | AY.12 | **AY.4** | 37.2 |
| P5 | B.1 | ND | **AY.7.1** | 42.5 |
| PV_19 | B.1.1.161 | ND | **B.1** | 35.5 |
| PV_8 | B.1 | ND | **B.1.1.28** | 39.8 |
| PV_17 | ND | ND | **B.1.2** | 39 |
| PV_18 | **A.5** | ND | **A.5** | 36.3 |
| PV_15 | B | ND | **B.1.1** | 37.3 |
| PV_16 | ND | ND | **B** | 39 |
| PV_20 | ND | ND | ND | 44.9 |

## DISCUSSION

Whole-genome sequencing (WGS) of SARS-CoV-2 has been performed extensively and provided essential information for public health. Molecular assays for COVID-19 detection and diagnosis are developed and updated based on SARS-CoV-2 sequence data. High-resolution data obtained by WGS made it possible to conduct precise epidemiological research for studying disease transmission and population dynamics. SARS-CoV-2 evolution has been closely monitored by WGS to identify the emergence and the spread of many variants of the virus. Furthermore, WGS played key roles in development of therapeutics and vaccines.

In this manuscript, two highly efficient SARS-CoV-2 sequencing protocols for Illumina sequencing platform were analyzed and summarized. Both COVIDSeq and SNAP protocols are amplicon-based SARS-CoV-2 genome sequencing protocols. RNA molecules contained in nasopharyngeal- and throat swab samples are a mixture of RNA molecules derived from the host and virus. After RNAs are isolated and converted into cDNA, cDNAs that contain SARS-CoV-2 genomic sequences are specifically selected and amplified by PCR-based target enrichment. COVIDSeq is performed using ARTIC network primers (https://github.com/artic-network/artic-ncov2019) to generate 98 amplicons (V1). The enriched PCR products were then fragmented and tagged by tagmentation. SNAP protocol generates 345 amplicons sized 116–255 bp for tagging without further fragmentation. The overlapping primers used in SNAP protocol cover 99.7% of the 29.9 kb viral genome.

Our results indicates that both COVIDSeq and SNAP kits generate high sequence coverage for SARS-CoV-2. We improved the yield by increasing input RNA concentration and using maximum number of PCR cycles. Digital PCR was able to provide better cutoff values for SARS-CoV-2 WGS than real-time PCR Ct values. Sequencing of all the samples with Ct ≤ 35 or >10.7 copies/μL by SNAP protocol or majority of the samples with Ct ≤30 or > 147 copies/μL by COVIDSeq achieved >95% genome coverage. There could be several contributing factors for the high recovery rate of SNAP protocol including using smaller size amplicons and normalizing the library load by normalase treatment prior to sequencing.

Since generating consensus FASTA was through reference mapping, combining FASTQ files generated from both protocols were not only possible but also provided several advantages. Though both COVIDSeq and SNAP kits use amplicon-based strategy, the regions that resulted in good sequence read from each protocol are different. In addition, this combining FASTQ strategy can provide effective tools to cope with sequencing of any emerging variants caused by amplicon loss due to novel mutations (17). Combining the FASTQ files increase the portions of the SARS-CoV-2 genomes covered by proper sequence read thereby improving the sequencing coverage outcome. By applying this, we were able to achieve >95% genome coverage for 113 samples and >99% genome coverage for 106 samples out of 121 samples which includes 42 samples of Ct 35∼45. Increased sequencing coverage made it possible to obtain accurate SARS-CoV-2 lineages.

Based on our experimental analysis we suggest the following for low viral load samples to achieve high sequencing coverage and accurate lineage information. 1. Increase RNA concentration by increasing sample loading and/or reducing elution volume; 2. Quantify viral load by dPCR; 3. For samples Ct>35, increase number of PCR cycles and apply normalase that comes with SNAP library kit; 4. Combine FASTQ files from COVIDseq and SNAP.

## Data Availability

All data produced in the present study are available upon reasonable request to the authors.

## Acknowledgements

This work was supported by a grant by VA SeqCURE which in turn received funding from the American Rescue Plan Act funds (Grant# N/A) with additional support from Central Texas Veterans Healthcare System (Temple, TX). The views expressed in this article are those of the authors and do not necessarily represent the views of the Department of Veteran Affairs or the funding agency.

We acknowledge CTVHCS clinical microbiology lab staff members; Mr. Shawn Sharp and Ms. Linda Wiley, Ms. Ivy Englett and Ms. Ma Rowena San Juan for organizing and testing clinical samples.

## Author contributions

Authors’ contributions: All authors have read and approved the final manuscript. All authors equally contributed to the data collection, analysis, and write up of the manuscript.

## FIGURE LEGENDS

**Table S1.** Sequence coverage obtained from SNAP, COVIDSeq, and combined FASTQ.

|  | Ct VALUE | % of non-N bases (SNAP) | % of non-N bases (COVIDSeq) | % of non-N bases (Combined) | Median Coverage (SNAP) | Median Coverage (COVIDSeq) | Median Coverage (Combined) |
| --- | --- | --- | --- | --- | --- | --- | --- |
| PV2_31 | 30 | 99.78% | 97.12% | 99.78% | 15039 | 2894 | 24759 |
| PV2_23 | 30 | 99.78% | 97.91% | 99.78% | 14116 | 3191 | 23370 |
| PV2_34 | 27 | 99.78% | 97.28% | 99.78% | 13988 | 2451 | 21806 |
| PV2_38 | 27 | 99.78% | 97.05% | 99.78% | 13713 | 3587 | 26200 |
| PV2_2 | 27 | 99.78% | 97.48% | 99.78% | 12759 | 2919 | 21708 |
| PV2_18 | 29 | 99.78% | 98.18% | 99.78% | 12746 | 3187 | 22526 |
| PV2_7 | 21 | 99.78% | 98.27% | 99.78% | 11628 | 3827 | 21590 |
| PV2_8 | 33 | 99.78% | 95.06% | 99.78% | 11375 | 2424 | 18804 |
| PV2_3 | 25 | 99.78% | 98.27% | 99.78% | 11141 | 3827 | 24483 |
| PV2_35 | 42 | 99.78% | 94.92% | 99.78% | 9493 | 1902 | 15326 |
| PV2_50 | 34 | 99.78% | 96.92% | 99.78% | 8948 | 1958 | 14717 |
| PV2_30 | 25 | 99.78% | 98.87% | 99.78% | 8333 | 4415 | 24025 |
| PV2_26 | 37 | 99.43% | 86.60% | 99.43% | 7181 | 761 | 9959 |
| PV2_93 | 19 | 99.74% | 98.97% | 99.78% | 7008 | 4354 | 21846 |
| PV2_76 | 27 | 99.78% | 96.57% | 99.78% | 5894 | 3578 | 17239 |
| PV2_25 | 35 | 99.15% | 69.45% | 99.46% | 5853 | 276 | 7055 |
| PV2_70 | 29 | 99.77% | 96.90% | 99.78% | 5852 | 2993 | 14584 |
| PV2_47 | 36 | 99.78% | 95.32% | 99.78% | 5691 | 1079 | 9210 |
| PV2_55 | 21 | 99.78% | 98.88% | 99.78% | 5429 | 3423 | 15591 |
| PV2_48 | 41 | 99.78% | 92.65% | 99.78% | 4784 | 1154 | 7891 |
| PV2_67 | 36 | 99.78% | 96.19% | 99.78% | 3868 | 2970 | 12870 |
| PV2_42 | 17 | 99.78% | 94.23% | 99.78% | 3806 | 1390 | 10980 |
| PV2_42 | 17 | 99.78% | 94.23% | 99.78% | 3806 | 1390 | 10980 |
| PV2_91 | 19 | 99.65% | 98.33% | 99.78% | 2913 | 5421 | 23437 |
| PV2_64 | 36 | 99.78% | 96.57% | 99.78% | 2429 | 2526 | 8991 |
| PV2_49 | 20 | 99.74% | 98.90% | 99.78% | 1966 | 4323 | 15410 |
| PV_11 | 40 | 98.97% | 70.33% | 99.61% | 1912 | 483 | 4072 |
| PV2_62 | 17 | 99.68% | 99.18% | 99.78% | 1732 | 2447 | 6335 |
| PV_12 | 41 | 97.98% | 84.03% | 99.09% | 1716 | 1691 | 6560 |
| PV2_89 | 42 | 99.78% | 97.48% | 99.78% | 1462 | 2627 | 8194 |
| PV2_83 | 31 | 98.06% | 69.13% | 99.46% | 1289 | 190 | 2319 |
| PV_6 | 43 | 98.38% | 78.29% | 99.39% | 1179 | 479 | 3024 |
| PV2_39 | 13 | 98.76% | 99.23% | 99.78% | 1004 | 5566 | 17117 |
| PV2_40 | 24 | 99.44% | 98.65% | 99.78% | 965 | 3201 | 13141 |
| PV2_82 | 34 | 96.52% | 76.03% | 98.94% | 736 | 358 | 1871 |
| PV_5 | 42 | 93.29% | 59.70% | 96.89% | 735 | 74 | 1416 |
| PV_13 | 16 | 99.32% | 99.78% | 99.78% | 721 | 8475 | 39413 |
| PV2_63 | 14 | 98.74% | 99.74% | 99.78% | 635 | 3085 | 7909 |
| PV_7 | 37 | 93.57% | 52.41% | 96.77% | 493 | 32 | 940 |
| PV_10 | 40 | 94.09% | 52.74% | 96.69% | 492 | 72 | 1335 |
| PV_4 | 37 | 95.49% | 75.25% | 98.83% | 482 | 225 | 1329 |
| PV2_58 | 37 | 99.23% | 90.81% | 99.76% | 427 | 1226 | 2603 |
| PV_2 | 38 | 90.71% | 79.89% | 98.65% | 345 | 154 | 891 |
| PV_8 | 40 | 83.56% | 30.62% | 90.54% | 311 | 5 | 534 |
| P5 | 42.5 | 82.68% | 44.35% | 90.38% | 287 | 14 | 408 |
| PV_3 | 45 | 85.46% | 54.12% | 92.46% | 241 | 87 | 851 |
| PV_1 | 37 | 85.73% | 74.13% | 96.81% | 231 | 75 | 619 |
| PV_15 | 37 | 62.31% |  | 66.78% | 164 |  | 181 |
| PV_18 | 36 | 56.71% | 0.157 | 64.72% | 147 | 3 | 179 |
| PV_19 | 36 | 60.77% | 39.84% | 78.96% | 140 | 11 | 244 |
| PV_17 | 39 | 45.51% | 0.2245 | 62.33% | 79 | 5 | 110 |
| PV_16 | 39 | 45.51% |  | 50.18% | 79 |  | 93 |

